# The Temporal Investigation of Multimodal Elements (TIME) Study: Protocol for an observational, longitudinal study to characterize the dynamic structure of molecular and digital data in healthy older adults

**DOI:** 10.64898/2026.05.14.26353203

**Authors:** James T. Yurkovich, Emma Glass, Noah Levine, Sophia Lee, Katheryn Ehlen, Eduardo Hernandez, Prabin Gharti, Ashen Fernando, Drew Witherington, Lance Pflieger, Jyotika Erram, Noa Rappaport, Alison Le, John C. Newman, Brianna Stubbs

## Abstract

**Background:** Biological systems exhibit dynamic patterns over multiple temporal scales—from minutes to months—that are poorly captured by conventional cross-sectional or low-frequency longitudinal studies. These patterns, including circadian and ultradian rhythms, may be critical determinants of health, resilience, and disease risk in aging. Existing longitudinal studies in older adults lack high-frequency, multimodal measurements that integrate molecular, physiological, and digital health data streams.

**Objectives:** The TIME Study aims to: (i) Characterize temporal patterns in molecular, physiological, and digital health measures in healthy older adults; (ii) determine how these patterns vary across biological domains and relate to each other; and (iii) assess how physiological systems respond to defined perturbations (oral glucose tolerance and maximal exercise).

**Methods:** TIME is a single-site, observational, longitudinal study enrolling up to 150 adults aged ≥ 55 years. Over an 11-week main phase, participants complete seven weekly low-frequency visits, two perturbation challenge visits, and two, two-day high-frequency sampling epochs. Biospecimens, clinical measures, cognitive and physical performance tests, and continuous digital health data are collected. Follow-up visits occur at 6 and 12 months.

**Expected Impact:** By integrating multimodal, temporally resolved data, TIME will provide a foundational dataset for understanding the role of biological rhythms in aging and inform future precision health strategies.

## 1 Introduction

The human body is a complex system that operates across multiple timescales, from the rapid firing of neurons to the slow, gradual changes of aging. These interconnected processes are often studied in isolation, leading to a fragmented understanding of human physiology^1,2^. As biomedical technology rapidly develops, there is a critical need to understand how these innovations can be best applied to human health^3^. One challenge is that many newer molecular profiling technologies (e.g., proteomics, metabolomics, lipidomics, methylation) have not been rigorously studied to understand how the time of day, fasting, or exercise impacts measurements^4–6^. This lack of understanding creates a barrier to translating these technologies into clinical practice, where they could provide valuable insights into health and disease^3,7^.

Concurrently, the landscape of patient monitoring is being transformed by digital health technologies (DHTs), which offer a scalable solution to the limitations of traditional, episodic clinical assessments ^8,9^. By enabling continuous, non-invasive data collection in real-world settings, wearables and mobile health platforms democratize access to high-frequency physiological tracking while reducing clinical costs and patient burden^10,11^. While these tools provide real-time resolution of biometric rhythms such as sleep patterns, activity, and resting heart rate, their full predictive utility is hindered by a disconnect from the patient’s underlying biochemical state. Despite parallel advancements in digital sensors and deep phenomic profiling, there remains a critical gap in rigorously integrating continuous digital health streams with deep biological measurements to holistically map the multi-scale temporal dynamics of human physiology^12^.

Here, we present the Temporal Investigation of Multimodal Elements (TIME) observational, longitudinal study that addresses this challenge by systematically mapping the biorhythms of the human phenome—the complete set of molecular and digital markers that define an individual’s biological state—over one year. The study’s overall goal is to provide a comprehensive understanding of how the human body changes over time, leading to the development of next-generation diagnostic tools and personalized health interventions. The TIME study’s primary hypothesis is that human biorhythms substantially impact molecular and physiological processes, thus directing the development of personalized and predictive health interventions. A key anticipated objective is to rigorously integrate continuous wearable metrics with high-resolution biochemical data, definitively bridging the gap between digital phenotypes and their underlying molecular drivers. The comprehensive temporal profiling provided here will reveal deep, novel insights into the dynamics of human physiology and result in a mapping of human biorhythms. This knowledge will pave the way for a future where healthcare is proactive, predictive, and tailored to the unique rhythms of each individual.

## 2 Methods

### 2.1 Study Design Overview

TIME is a single-site, observational, longitudinal study designed to characterize temporal biological patterns in healthy older adults. The study is primarily discovery-driven, recognizing that there is currently insufficient data to form specific hypotheses regarding time-of-day or week-to-week dependence for many omic endpoints (e.g., proteomics, metabolomics) in older adults.

Over a 2.5-year enrollment period, the TIME study will capture a wide range of time points and naturalistic conditions (Figure 1). Seasonal variation will occur and is not explicitly controlled for; instead, it is documented and treated as a feature of the dataset, providing an additional axis of variability to inform future targeted investigations.

**Figure 1:**
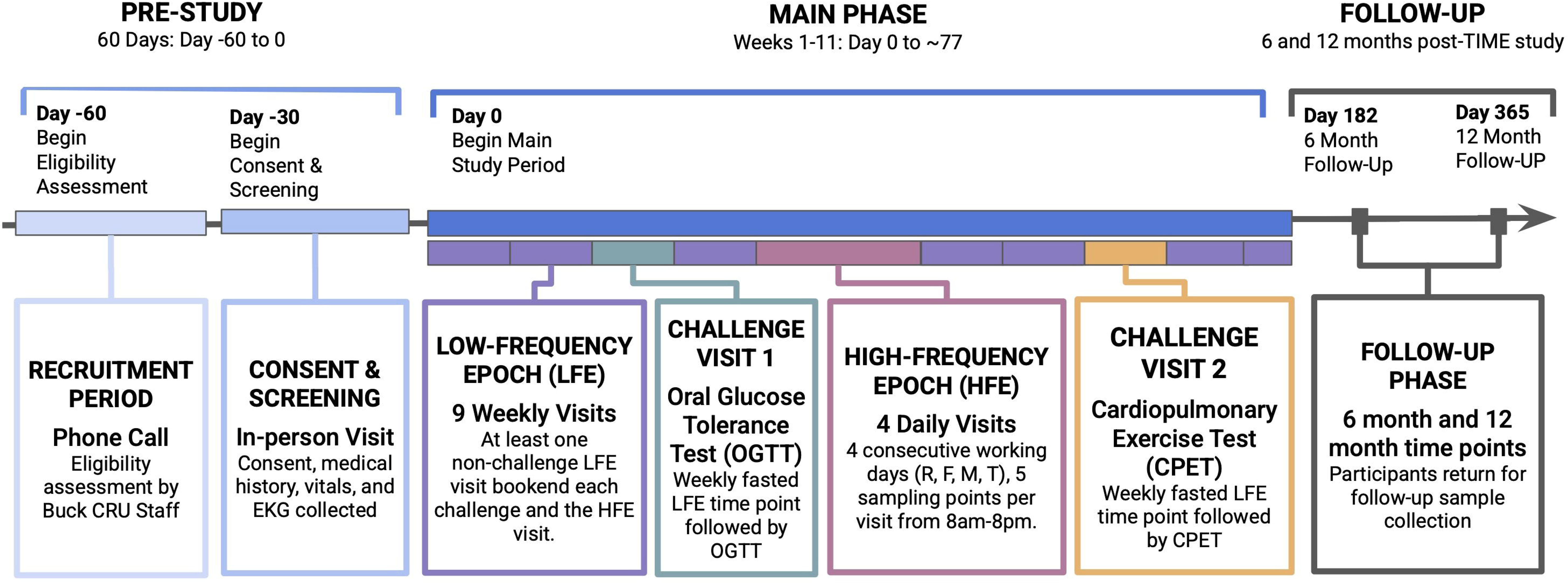
Overview of the TIME Study. At each week’s visit, blood, urine, saliva, stool, microbiome swabs are collected, as well as vitals, cognitive testing, physical function testing, retinal scans, and facial imaging. Oura Rings, WHOOP Bands, Stelo by Dexcom CGMs, and Withings Body Pro 2 scales are used throughout the study.

Participants will be studied in detail for the main 11-week period, and then a follow-up period will see the collection of biospecimens and measurements 6- and 12-months after enrollment. Digital health data will be collected throughout the main and follow-up periods. The main 11-week phase involves 7 low-frequency epoch (LFE, single fasted biospecimen collection and limited other measures), 2 challenge visits (including the normal LFE sampling), and 4 days of high-frequency epoch (HFE, 12 hours of regular biospecimen collection and comprehensive measures) on a consecutive Thursday, Friday, Monday and Tuesday. These timepoints were chosen to allow both week-to-week and intra-day assessment of biorhythms. The choice to do HF sampling at the beginning and end of the working week rather than across a week allows us to examine the impact of changing behaviors such as sleep, diet, or exercise over the weekend. We also decided to capture data from two consecutive days, twice, so that each individual can act as their own biological control across days.

On two of the weeks, participants will complete challenges, an oral glucose tolerance test, and a maximal exercise test; these will capture the response of the body to perturbations. These challenges were chosen as they are highly studied in the literature^13–16^, are known to produce large changes in usually stable biomarkers and provide outcome data (i.e., insulin resistance, VO_2_ max) that can be related directly to health (mortality, morbidity) and function. These two challenges also stress test different systems of the body—cardiovascular for exercise testing, and metabolic for the glucose tolerance test—further providing an opportunity to profile and characterize the participants’ physiology.

By capturing high-frequency (daily), low-frequency (weekly), challenge and follow-up data, the study will create a foundational dataset that can be used to bridge the gap between discrete molecular measurements and near-continuous digital health data.

The study protocol described here has been approved by Advarra Institutional Review Board (current version 1.0 approved on 7^th^ April 2025, IRB reference: Pro00085349), any further amendments will be approved by the IRB. Participants will also be informed of any protocol modifications affecting their study procedures and will provide updated consent (initials and date on the relevant ICF section) prior to undergoing any modified procedure. The study is registered with clincaltrials.gov (NCT07107386), all items from the WHO Trial Registration Dataset are included in Supplementary Table 1. Enrollment began on June 15^th^ 2025 and is expected to run until June 2027. Here we present details of the study protocol according to SPIRIT 2025 guidelines (Supplementary information).

### 2.2 Study Setting

The TIME Study is conducted by investigators and clinical research staff at The Buck Institute for Research on Aging, a non-profit research institution based in Northern California, USA, in partnership with Phenome Health, a Seattle-based nonprofit. The majority of participants are expected to be residents of Northern California.

### 2.3 Study Population

Participants in TIME (up to n = 150) will be adults at or over the age of 55 years in stable health. We aim to recruit an even number of male and female participants and a population representative of the demographics surrounding the clinical site. The choice to focus on adults over the age of 55 years was intended to reduce variability. Firstly, the human phenome is known to change across the lifespan^17^, narrowing the enrollment window reduces natural variability occurring with age^18^. Secondly, in females the menstrual cycle and the menopause transition are known to cause increased variability on both a micro and macro level^19,20^, and so recruiting a population without a menstrual cycle and likely to be post-menopause removes this potentially large source of variability. However, we strongly believe that a future study should apply the same dense phenomic sampling approach to younger adults, especially women ages 18-45 to examine changes across all phases of the menstrual cycle.

Eligibility criteria are intentionally broad to capture a range of health statuses representative of the older adult population, while ensuring safety for all study procedures. Stable chronic diseases are not automatically exclusionary unless judged uncontrolled or clinically active by the study medical officer. As this study involves frequent blood collection, participants with anemia, coagulopathies, recent blood donation, and poor venous access are excluded. Notably, participants must reside in the Pacific or Mountain time zone, may not undertake travel crossing more than two time zones during the main 11-week study, and may not be shift workers; these requirements aim to reduce possible effects of time zones on circadian biology. Similarly, we exclude participants using many sleep medications more than once per week in the three months prior to the initial visit. Finally, as we plan to collect many types of microbiome samples, we exclude any participant with antibiotic use within 3 months of the initial visit. Other medications and supplements are permitted if use has been stable for at least three months before the initial visits. These are recorded and may in fact be of interest during our multiomic analysis. If a participant is required to start a new medication, elects to start a new supplement, or has a new, non-life-threatening diagnosis during the study this is not exclusionary and will be recorded and examined during analysis. Full inclusion and exclusion criteria are found in Table 1.

**Table 1:**
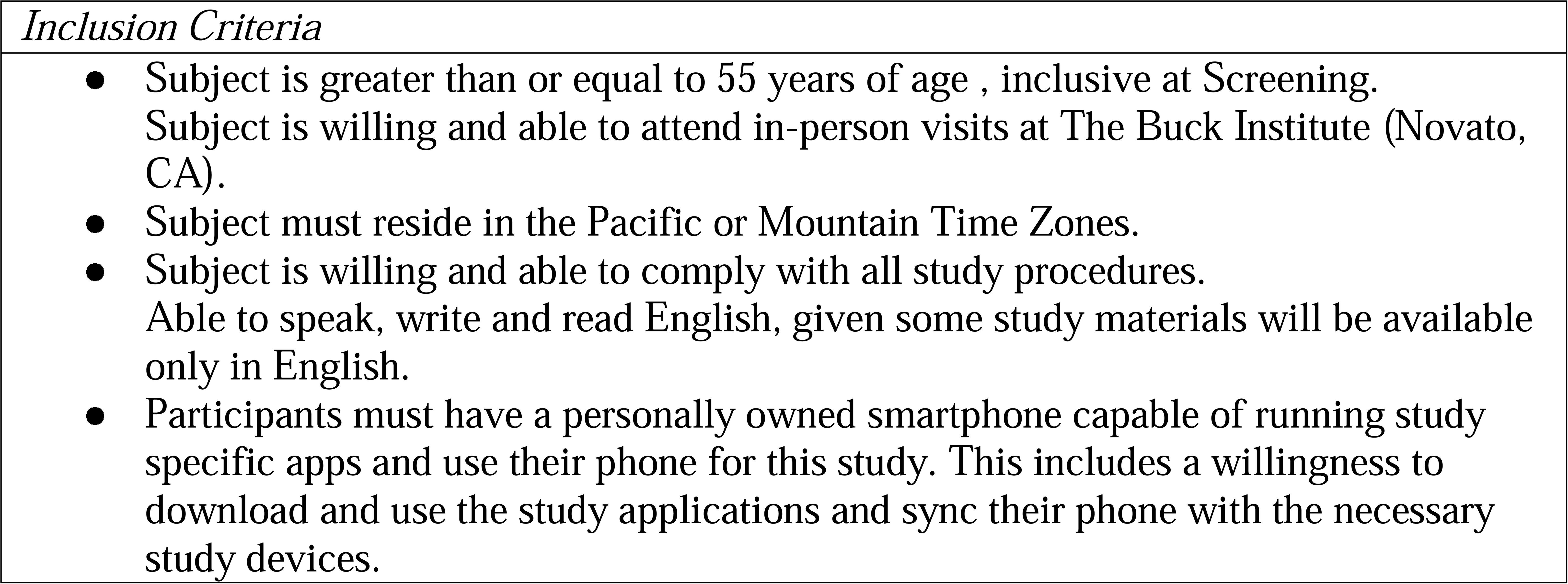
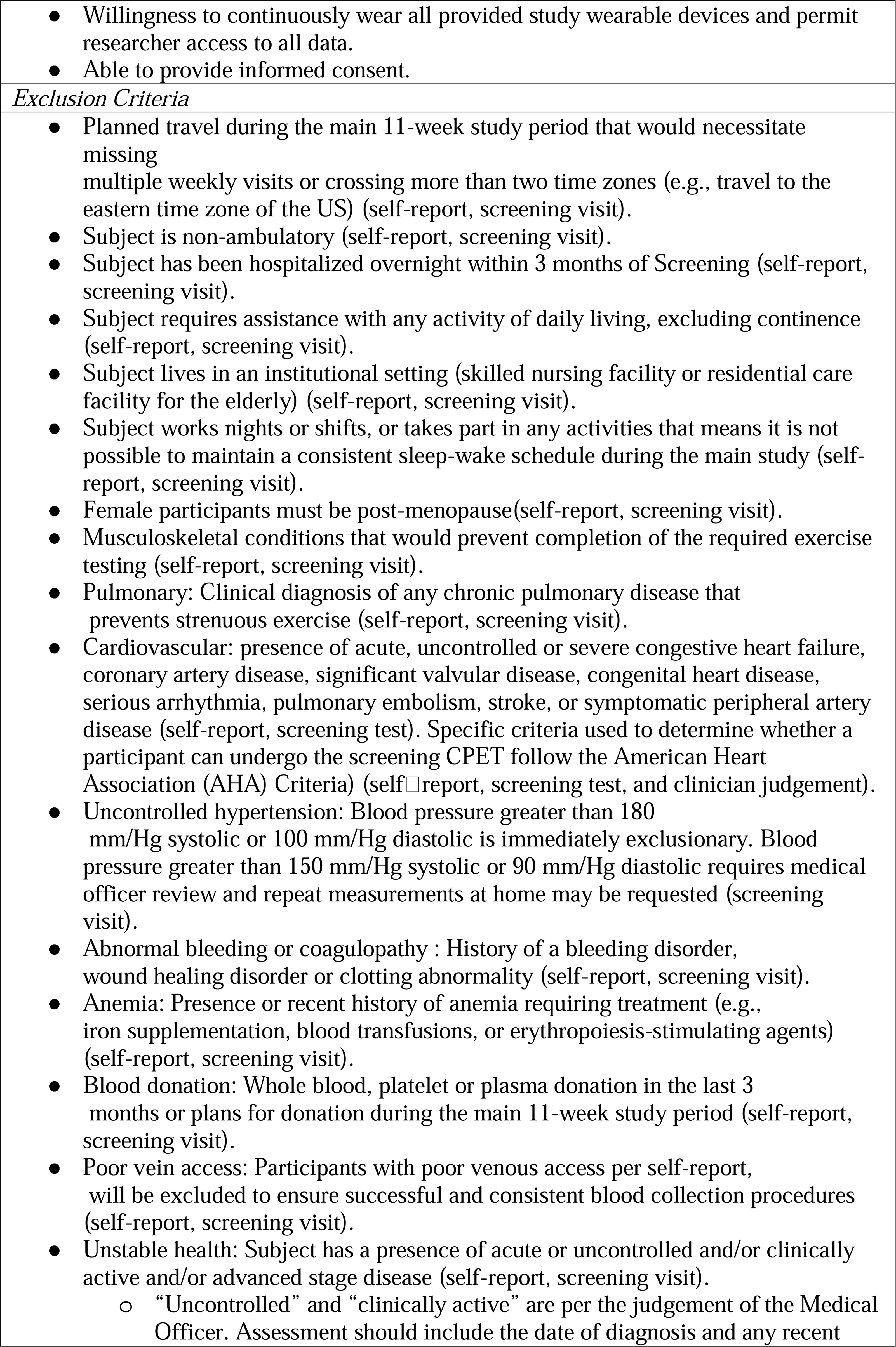
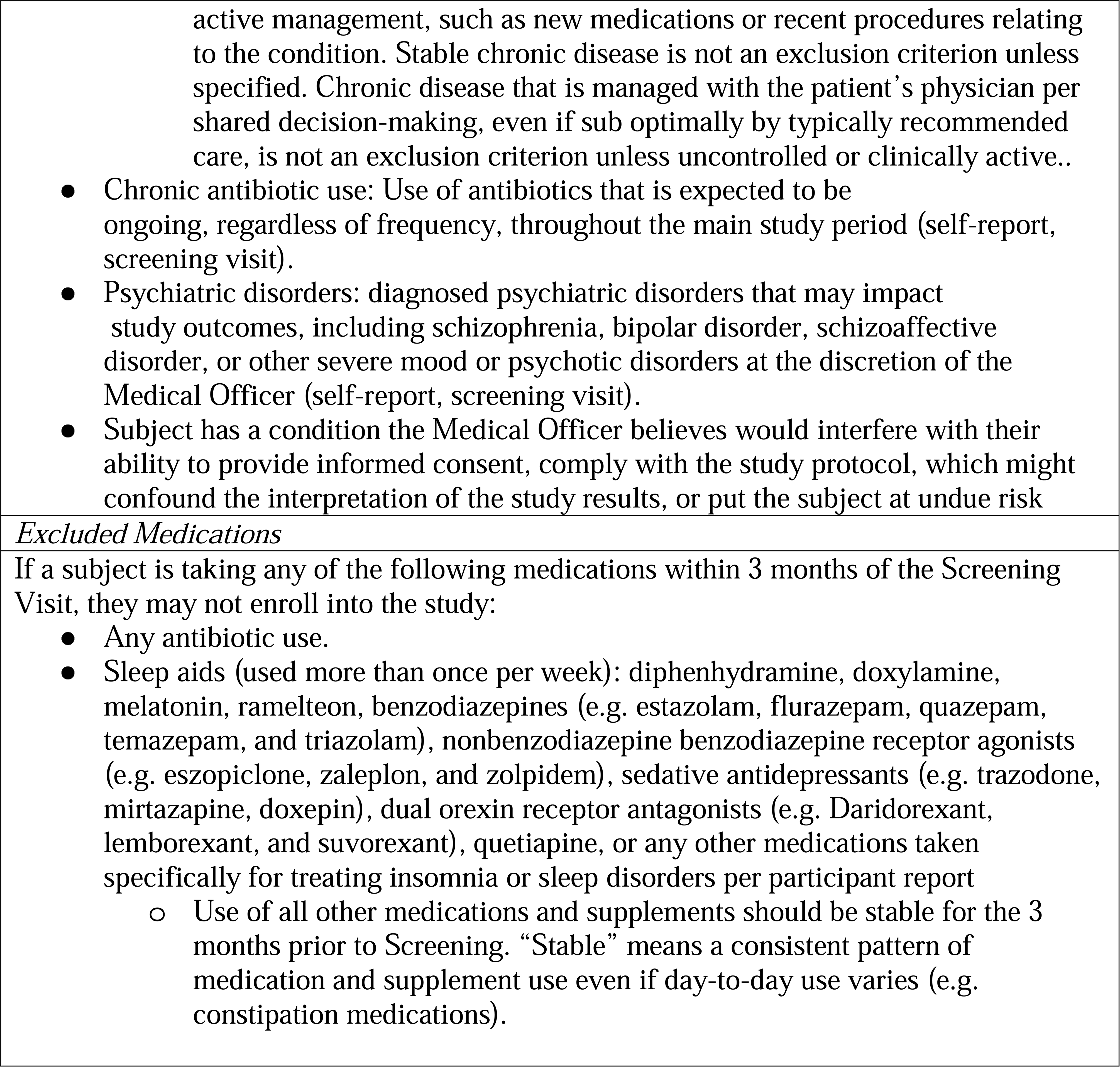
Inclusion/Exclusion Criteria.

### 2.4 Study Objective

The TIME study’s primary objective is to comprehensively map human biorhythms by characterizing the dynamic interplay of molecular and digital health markers across daily and weekly timescales. This will be achieved through the analysis of high-frequency molecular data, including proteomics, metabolomics, and lipidomics, alongside continuous physiological data streams from wearable sensors. Despite the rapid innovation in biological measurement technologies, the field has not identified a minimum viable dataset required to accurately characterize an individual’s unique, dynamic biological state. It remains undetermined which specific combinations of variables or datatypes—whether continuous digital phenotypes or discrete multi-omic signatures—serve as the most robust, actionable biomarkers. By systematically capturing these dense, multimodal data streams in tandem, the TIME study aims to construct a “biological Rosetta Stone”. This translational framework will map the complex correlations across diverse biological domains, allowing us to decipher how easily accessible, non-invasive wearable metrics reflect deep underlying biochemical fluctuations. Ultimately, establishing this biological Rosetta Stone will enable us to distill the vast human phenome down to an optimal, minimal viable dataset, identifying biological and digital biomarkers necessary to drive scalable precision health interventions.

### 2.5 Study Data

Data collection will occur from a range of biological samples, digital devices, participant reported measures, and clinical assessments. A Data Monitoring Committee is not required for this study because it is a single-site, observational study with minimal risk to participants. Study procedures are limited to standard clinical assessments, biospecimen collection, and non-invasive monitoring. The study team will internally oversee data collection and participant safety according to standard operating procedures. There are, of course, a multitude of functional, clinical, digital, and molecular technologies that can be used to profile human physiology. We identified two major considerations when choosing the technologies to be used in the TIME Study:

1. <u>Data interoperability</u>: To maximize the translational impact and utility of the TIME dataset, we prioritized analytical platforms that ensure seamless cross-compatibility with major, population-scale cohorts^21–26^. By selecting established, industry-standard technologies—such as the Olink platform for global proteomics and Metabolon for global metabolomics—we deliberately align our molecular readouts with the foundational protocols employed by massive longitudinal initiatives like the UK Biobank^22^ and the NIH *All of Us* Research Program^21^. This strategic harmonization enables researchers to leverage vast external reference datasets to contextualize the high-frequency temporal dynamics and personalized biorhythms mapped within our study.
2. <u>Future-proofing</u>: Recognizing the rapid evolution of multi-omic profiling and biomarker detection technologies, a core priority of the study design is the long-term future-proofing of our physical biorepository. We have instituted rigorous, standardized protocols for the high-volume collection, comprehensive processing, and long-term cryogenic banking of diverse blood fractions (e.g., plasma, serum, peripheral blood mononuclear cells). By establishing a high-quality physical archive alongside the immediate digital and molecular datasets, we ensure that as novel, higher-resolution analytical technologies inevitably emerge, these uniquely valuable, temporally dense longitudinal samples can be retrospectively analyzed^27^. This biobanking strategy maximizes the scientific yield of the study and guarantees that the TIME cohort remains an enduring, adaptable resource for future discoveries in predictive health.

#### 2.5.1 Digital Health Technologies

DHTs serve as the foundation of the TIME study’s longitudinal architecture, providing the continuous physiological context necessary to accurately interpret episodic clinical datatypes. By passively capturing a high-frequency stream of biometric and behavioral data in a natural, day-to-day setting, these tools establish a robust, real-world ground truth of each participant’s baseline state and daily lifestyle. Crucially, the deployment of advanced multimodal sensors—such as continuous glucose monitors, smart scales, and wearable devices—links the study’s low-frequency biological sampling time points. This dense digital phenotyping prevents the loss of vital dynamic information between physical clinic visits, allowing us to continuously monitor physiological strain, recovery, and biophysical fluctuations. Ultimately, anchoring deep, multi-omic molecular profiling to this continuous digital framework is what enables the accurate mapping of multi-scale temporal biorhythms. A summary of the specific DHTs deployed, alongside all current data sources and expected endpoints, is detailed in Table 2.

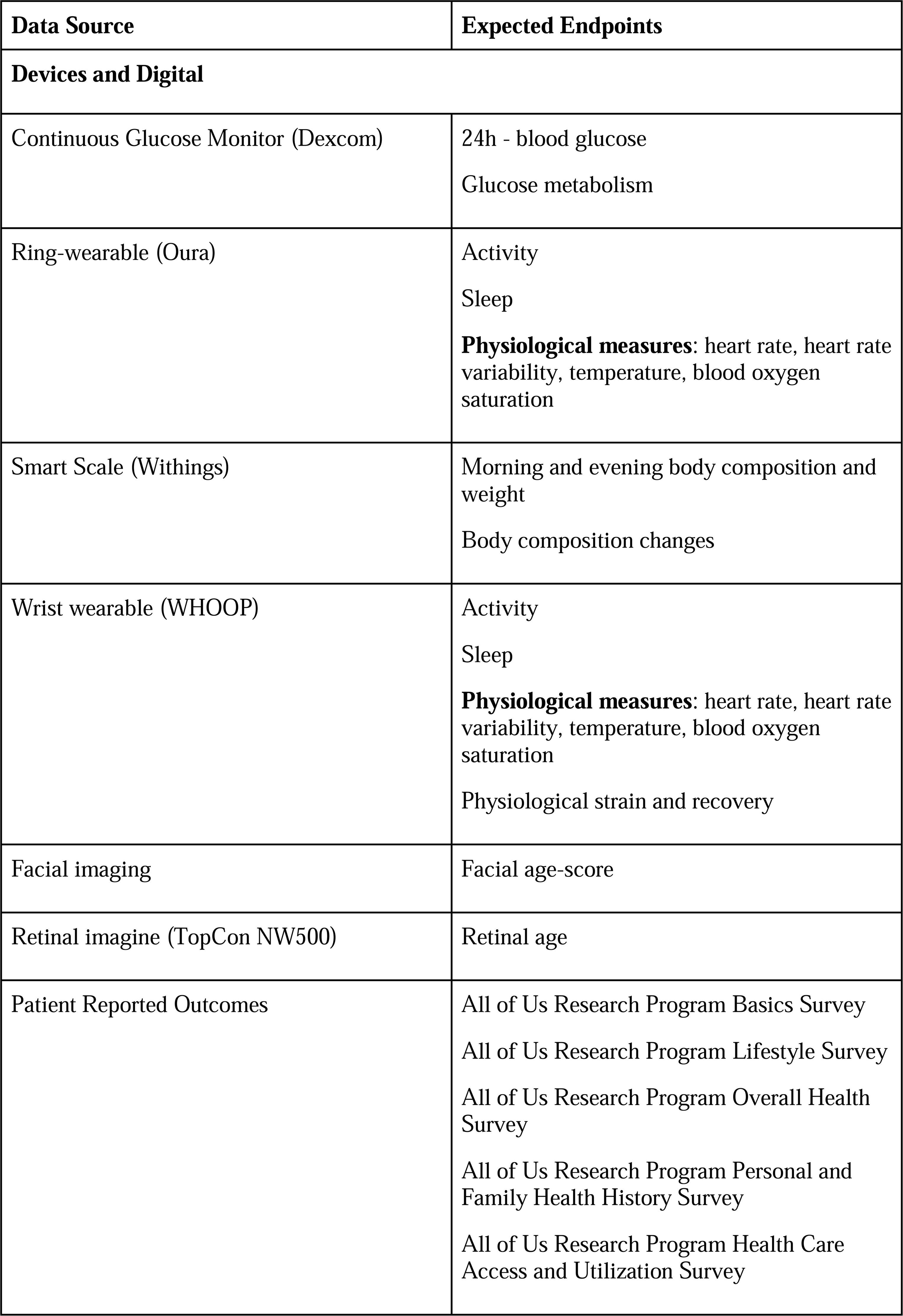
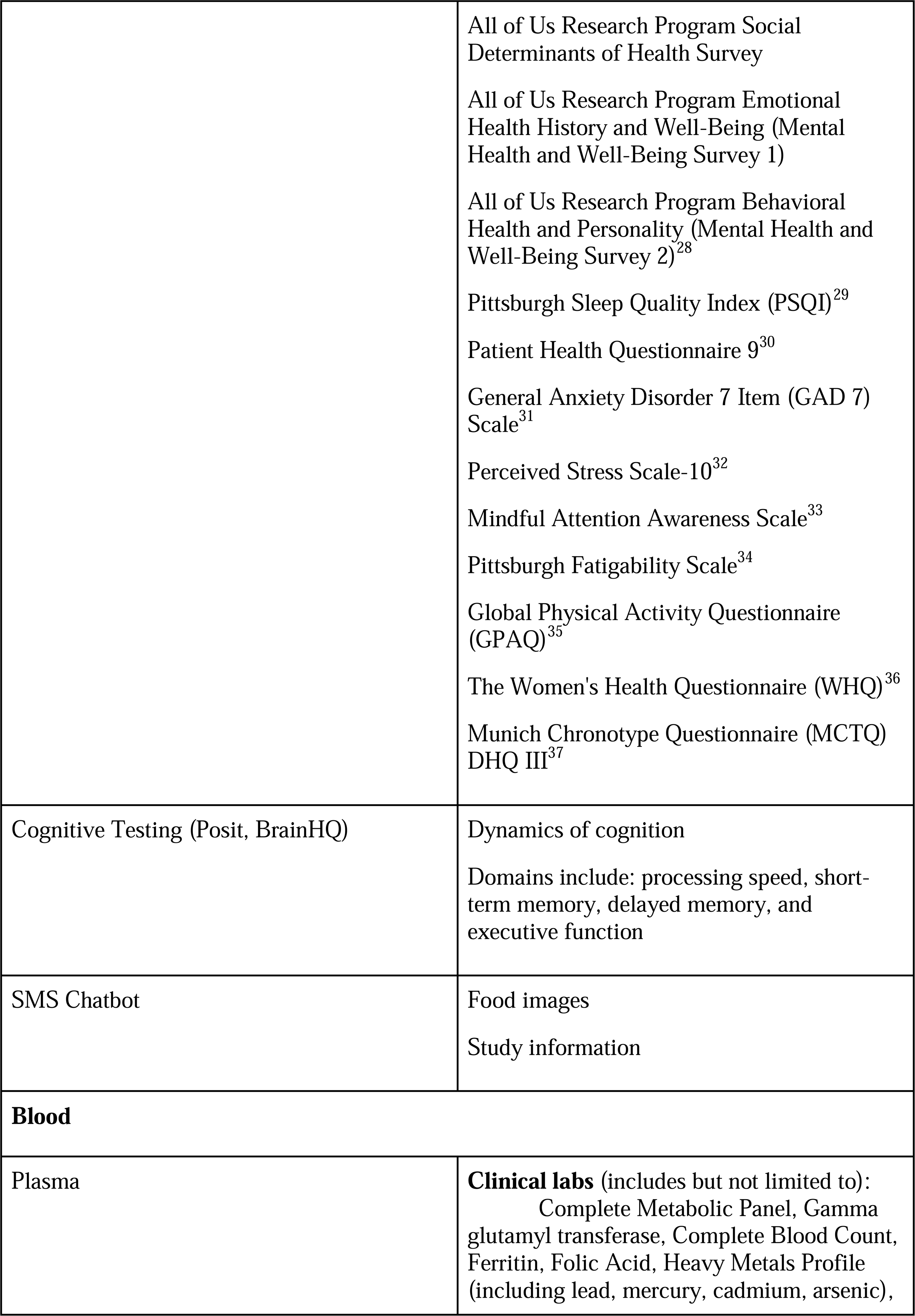
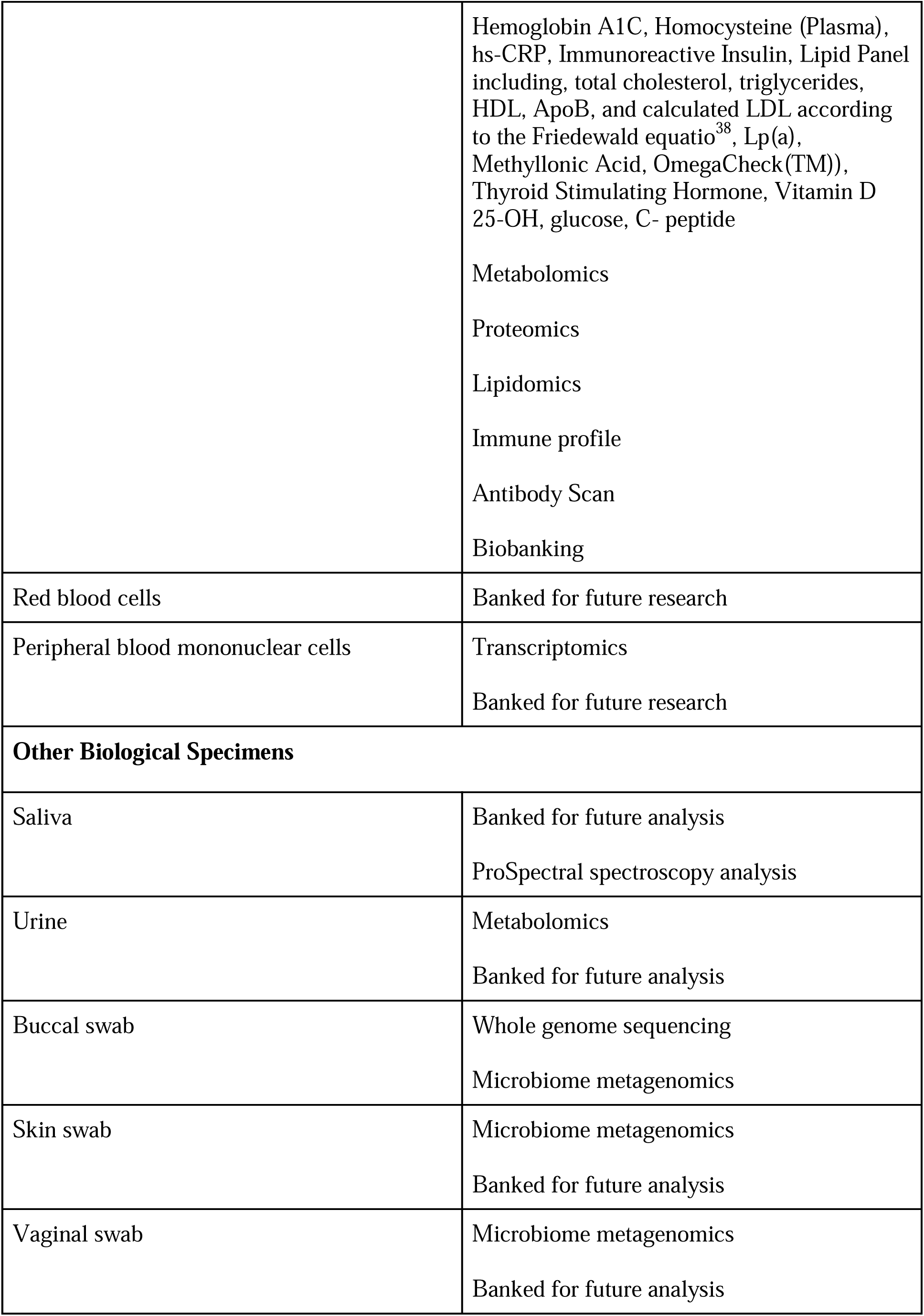
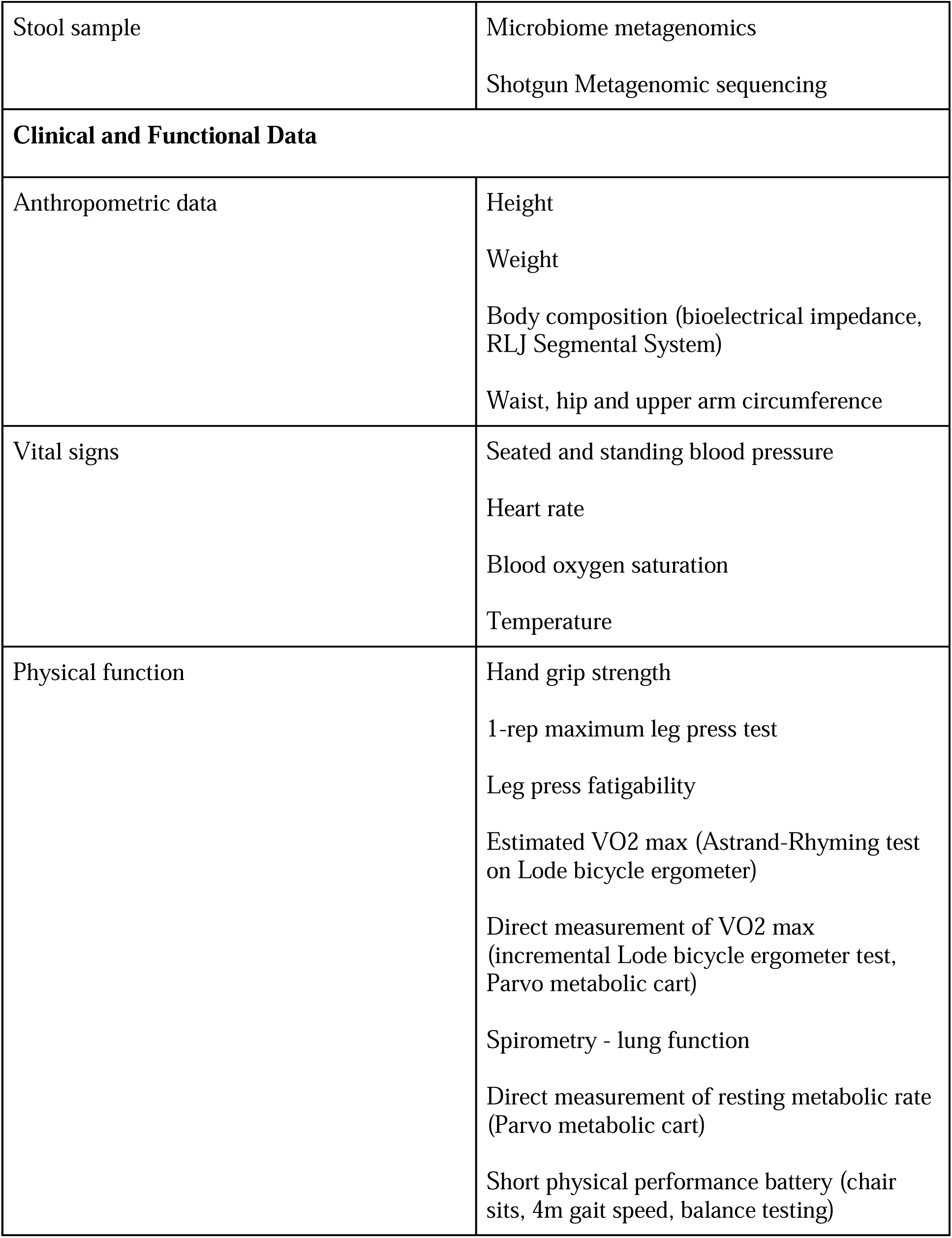

| Data Source | Expected Endpoints |
| --- | --- |
| **Devices and Digital** |  |
| Continuous Glucose Monitor (Dexcom) | 24h - blood glucose<br>Glucose metabolism |
| Ring-wearable (Oura) | Activity<br>Sleep<br>**Physiological measures:** heart rate, heart rate variability, temperature, blood oxygen saturation |
| Smart Scale (Withings) | Morning and evening body composition and weight<br>Body composition changes |
| Wrist wearable (WHOOP) | Activity<br>Sleep<br>**Physiological measures:** heart rate, heart rate variability, temperature, blood oxygen saturation<br>Physiological strain and recovery |
| Facial imaging | Facial age-score |
| Retinal imagine (TopCon NW500) | Retinal age |
| Patient Reported Outcomes | All of Us Research Program Basics Survey<br>All of Us Research Program Lifestyle Survey<br>All of Us Research Program Overall Health Survey<br>All of Us Research Program Personal and Family Health History Survey<br>All of Us Research Program Health Care Access and Utilization Survey |
|  | All of Us Research Program Social Determinants of Health Survey<br>All of Us Research Program Emotional Health History and Well-Being (Mental Health and Well-Being Survey 1)<br>All of Us Research Program Behavioral Health and Personality (Mental Health and Well-Being Survey 2)<sup>28</sup><br>Pittsburgh Sleep Quality Index (PSQI)<sup>29</sup><br>Patient Health Questionnaire 9<sup>30</sup><br>General Anxiety Disorder 7 Item (GAD 7) Scale<sup>31</sup><br>Perceived Stress Scale-10<sup>32</sup><br>Mindful Attention Awareness Scale<sup>33</sup><br>Pittsburgh Fatigability Scale<sup>34</sup><br>Global Physical Activity Questionnaire (GPAQ)<sup>35</sup><br>The Women's Health Questionnaire (WHQ)<sup>36</sup><br>Munich Chronotype Questionnaire (MCTQ) DHQ III<sup>37</sup> |
| Cognitive Testing (Posit, BrainHQ) | Dynamics of cognition<br>Domains include: processing speed, short-term memory, delayed memory, and executive function |
| SMS Chatbot | Food images<br>Study information |
| **Blood** |  |
| Plasma | **Clinical labs** (includes but not limited to):<br>Complete Metabolic Panel, Gamma glutamyl transferase, Complete Blood Count, Ferritin, Folic Acid, Heavy Metals Profile (including lead, mercury, cadmium, arsenic), |
|  | Hemoglobin A1C, Homocysteine (Plasma), hs-CRP, Immunoreactive Insulin, Lipid Panel including, total cholesterol, triglycerides, HDL, ApoB, and calculated LDL according to the Friedewald equation<sup>38</sup>, Lp(a), Methyllonic Acid, OmegaCheck(TM)), Thyroid Stimulating Hormone, Vitamin D 25-OH, glucose, C- peptide<br>Metabolomics<br>Proteomics<br>Lipidomics<br>Immune profile<br>Antibody Scan<br>Biobanking |
| Red blood cells | Banked for future research |
| Peripheral blood mononuclear cells | Transcriptomics<br>Banked for future research |
| **Other Biological Specimens** |  |
| Saliva | Banked for future analysis<br>ProSpectral spectroscopy analysis |
| Urine | Metabolomics<br>Banked for future analysis |
| Buccal swab | Whole genome sequencing<br>Microbiome metagenomics |
| Skin swab | Microbiome metagenomics<br>Banked for future analysis |
| Vaginal swab | Microbiome metagenomics<br>Banked for future analysis |
| Stool sample | Microbiome metagenomics<br>Shotgun Metagenomic sequencing |
| **Clinical and Functional Data** |  |
| Anthropometric data | Height<br>Weight<br>Body composition (bioelectrical impedance, RLJ Segmental System)<br>Waist, hip and upper arm circumference |
| Vital signs | Seated and standing blood pressure<br>Heart rate<br>Blood oxygen saturation<br>Temperature |
| Physical function | Hand grip strength<br>1-rep maximum leg press test<br>Leg press fatigability<br>Estimated VO2 max (Astrand-Rhyming test on Lode bicycle ergometer)<br>Direct measurement of VO2 max (incremental Lode bicycle ergometer test, Parvo metabolic cart)<br>Spirometry - lung function<br>Direct measurement of resting metabolic rate (Parvo metabolic cart)<br>Short physical performance battery (chair sits, 4m gait speed, balance testing) |

### 2.6 Participant Timeline

#### 2.6.1 Recruitment

Participants will be recruited using advertisements via email, website banners, social media, flyers, community outreach and partnerships, and referrals. Recruitment materials will be designed to provide clear and concise information about the study, including its purpose, eligibility criteria, compensation, and contact information for interested individuals. Potential participants will be identified based on the study’s inclusion and exclusion criteria and approached through in-person interactions at community and private events, phone calls, emails and mailed invitations.

Efforts will be made to ensure recruitment reflects the population of the San Francisco Bay Area, with targeted outreach to senior community centers, independent living communities, clubs, gyms, religious centers, and academic institutions to enhance the generalizability of study findings.

#### 2.6.2 Telephone screening

During the telephone screen, subjects will be provided with a summary of the study and assessed for initial eligibility.

#### 2.6.3 In-person visits

For all in-person visits, participants must meet the following pre-test criteria: fasting ≥ 10 hours, no alcohol ≥ 10 hours, no strenuous exercise ≥ 10 hours, no psychoactive cannabis products ≥ 10 hours. Participants will be queried on arrival to confirm compliance. At every visit, participants will be queried on their use of any new medications and supplement, or any changes to their health. Participants will also be queried to ensure compliance with study protocol, and will be reminded of study instructions.

As the main phase of the protocol is strenuous, requiring weekly in-person visits for 11 weeks, we allow some flexibility in timing to facilitate participant retention. One LFE in-person visit must occur per working week (Monday – Friday) during the 11-week main period; there will be an allowable window of minimum four days and a maximum of nine days between all study visits. Similarly, if nine visits cannot be completed within 11 weeks, it is allowable to take up to 15 weeks to complete all required visits. LFE visits do not take place during the two working weeks when HFE visits are scheduled. In addition, we plan to flexibly schedule the two challenge visits and the four-day high frequency sampling block. A final relevant retention strategy will be to pay stipends in a staged manner, increasing in value the further a participant progresses through the study.

There must be a non-challenge, LFE weekly visit before each of the challenge visits and before the HFE sampling block. The oral glucose tolerance challenge occurs first (between Week 2-5), followed by the HFE sampling block (Weeks 4-8) and lastly the maximal exercise test (Weeks 6-10).

Given the study’s core focus on multi-scale temporal dynamics, all biospecimen collections will be strictly time-stamped upon acquisition, and strict adherence to standardized, time-gated processing windows—from initial draw to final preservation—will be enforced to eliminate pre-analytical temporal confounding. A study timeline in the SPIRIT Figure format is shown in the Supplemental Information.

#### 2.6.4 Screening Visit (Visit 1)

Staff will review the informed consent form with potential participants, provide time for questions, and confirm understanding before obtaining participant signature. Consent will be conducted in person in a private setting in accordance with Good Clinical Practice guidelines. The consent process will include options for broad data and biospecimen sharing for future research, as applicable. All study procedures are considered minimal risk. Blood draws, exercise testing, and glucose tolerance testing are standard procedures; adverse events are monitored per protocol.

Participants will provide written informed consent and a full medical history, medication and supplement use, demographic data, vital signs and a resting EKG will be obtained. These data will be reviewed by a study medical officer who will make final decisions on eligibility and enrollment into the study. No additional ancillary or post-trial care or compensation for research-related injury will be provided. Participants will be informed of this limitation during the informed consent process. Participants will be advised to seek medical care through their own healthcare providers in the event of any adverse health events arising during the study.

### 2.7 Main Phase (11 weeks)

#### 2.7.1 Low Frequency Visits (9 visits over 11 weeks)

At a morning visit, a single LFE data collection time point will take place (∼0800H). This consists of a vital sign measurement, venous blood sample, a urine sample, a retinal scan, a saliva sample, a buccal swab, a skin swab, a vaginal swab (female subjects only), cognitive testing and grip strength testing. Subjects will be given at-home kits for collection of a stool sample within the 3 days prior of all LFE visits.

At the first and last LFE visits additional procedures will be performed. This will include anthropometric measures, a subset of the self-reported measures, resting metabolic rate measurement , and set-up and distribution of study devices (first LFE visit only). At the LFE visit prior to both challenge visits and the HFE visits, a continuous glucose monitor will be applied to the upper arm, and participants will be given standard meals to consume prior to the challenge or HFE visits.

#### 2.7.2 At-home Procedures

Participants will be asked to wear and sync study wearable devices throughout the 12-month study, and to record their weight twice daily using the Smart Scale (Withings Body Pro 2). To incentivize participation and increase retention, participants who complete the main study period may keep the study devices. During the main 11 week study period participants will be asked provide to routine data tags about activities (e.g., exercising, napping) and other self-measurements (e.g., blood pressure) via SMS chatbot, additionally during the three two-week periods where a CGM is worn, participants will be asked to photograph all food consumed and share via SMS Chatbot. If a CGM fails or falls off before the 15-day window, it is replaced if feasible.

#### 2.7.3 SMS Chatbot

The current state of the art in behavior tracking is transitioning away from high-burden, app-based logging toward low-friction, conversational interfaces. By utilizing natural language processing and computer vision directly within native messaging platforms, modern ecological momentary assessment (EMA) tools significantly reduce recall bias and participant fatigue compared to traditional dietary and lifestyle diarie^39–42^. The SMS Chatbot, a next-generation AI-backed system, is designed to support the TIME Study by collecting data in a natural, conversational manner. Its primary function is to engage with participants via SMS texting (i.e., no additional mobile application required) to collect additional data and information. This is achieved through two types of interactions: user-initiated messages, such as sending photos of food; and SMS Chatbot-initiated messages, which are scheduled to collect information at specific times, such as around meals or after a workout. These data are timestamped and correlated with other data collected from the participants’ wearables.

The SMS Chatbot is governed by strict guardrails, ensuring the accuracy and completeness of its responses. Its functionality is intentionally limited to a predefined set of tasks, including wearable integration, daily habit tracking, food and nutrition logging, and user data retrieval. The knowledgebase is derived from a curated selection of peer-reviewed literature and books, and it is explicitly designed to not provide medical advice. To further ensure security and prevent manipulation, the SMS Chatbot undergoes rigorous daily testing. The goal of this system is to increase retention and engagement of study participants by answering questions and facilitating the collection of additional data from questionnaires, surveys, and daily occurrences.

#### 2.7.4 Study Meals

Study participants will be consuming a total of 25 standardized study meals immediately before and during the challenges. During in-clinic HFE visits, meals are provided at approximately 0845, 1200, and 1800H. Meals are designed to be congruent with a Mediterranean dietary pattern in their macronutrient and micronutrient composition. Participants select from a menu of 11 pre-made options, with omnivore, gluten-free, and vegetarian selections available to accommodate a range of dietary preferences. Total daily energy expenditure is estimated for each participant using the Mifflin-St Jeor equatio^43^ with a sedentary activity factor, reflecting the limited physical activity expected during in-clinic study days. Supplemental nutrition shake(s) are provided alongside the study meals as needed to match total energy intake to estimated expenditure, maintaining an energy-neutral balance throughout the study meal periods. Meals are delivered frozen and reheated on site; for at-home meal periods, participants receive frozen take-home meals. Compliance with meal and shake consumption is assessed by photographing each item on a scale before and after eating to quantify intake by weight. During at-home meal periods, participants are free to consume their provided meals at times of their choosing. Samples of the meals will be saved for future molecular analysis (e.g., using mass spectrometry) to perform integrative analyses with corresponding molecular data being generated from biospecimens.

#### 2.7.5 Challenge Visit: Oral Glucose Tolerance Test (between Weeks 2 – 5)

At the start of the visit, an IV cannula will be inserted and the LFE fasting blood sample will be collected following completion of the other LFE measures, directly before the OGTT is initiated. Participants will be asked to consume a drink containing a standardized amount (75 grams) of glucose (Glucola) within 5 minutes. Venous blood sample collection will occur at 10, 20, 30, 60, 90, 120 and 180 (+/-5) minutes after administration of the oral glucose bolus (Figure 2).

**Figure 2.**
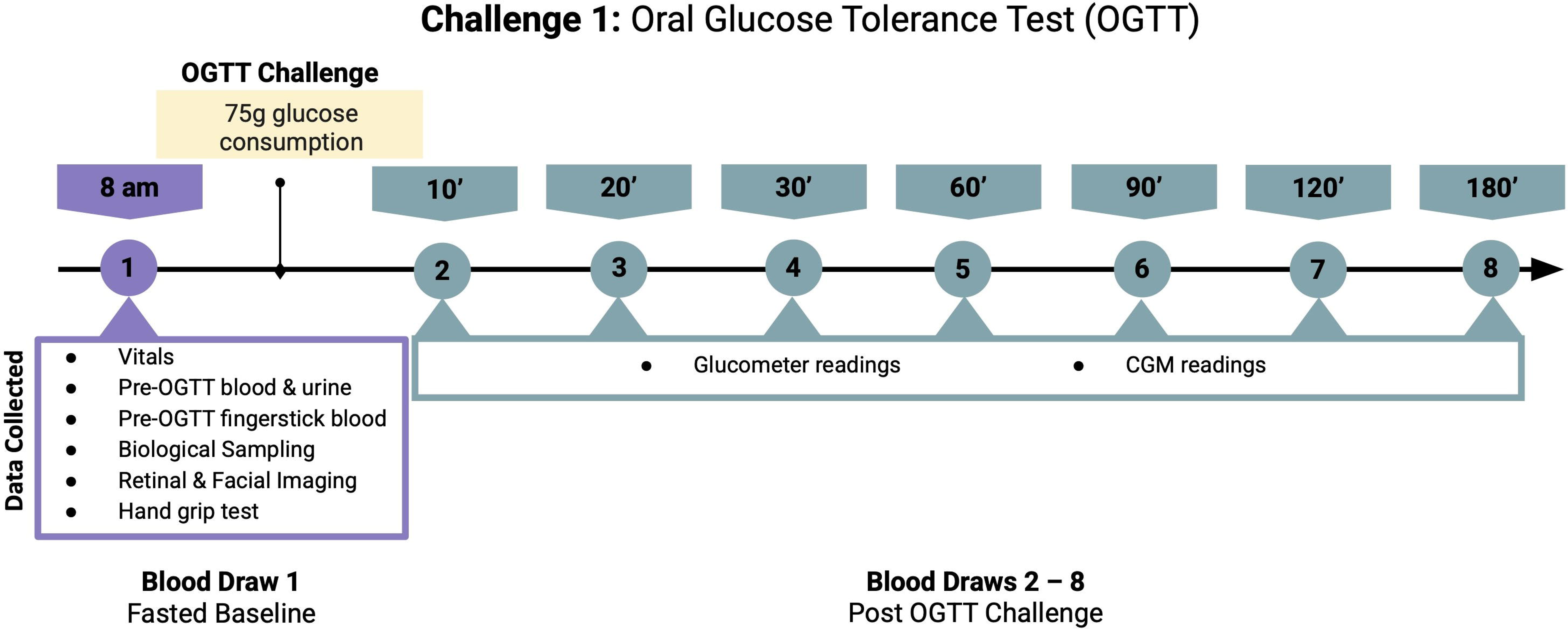
Challenge 1: Oral Glucose Tolerance Test Sampling Schedule

#### 2.7.6 High Frequency Sampling Block (between weeks 4 – 8)

Four near-identical in-person HFE visits occur over ∼12 hours (0800 - 2000H) on a Thursday (Visit 1), Friday (Visit 2), Monday (Visit 3), Tuesday (Visit 4). Whilst we considered that this schedule might alter a participant’s regular sleep-wake schedule, data from the wearable devices will allow us to account for the possible interaction between an individual’s circadian timing and sample timing, and a set schedule mimics the nature of many other studies and clinical procedures that require fasting, as well as increasing operational feasibility for the clinical team.

Data will be collected at five time points: 0800, 1100, 1400, 1700 and 2000H. Data collected at each time point varies, but includes blood (from an IV cannula), urine, saliva, retinal imaging, facial imaging, cognitive function, grip strength, buccal swabs, vaginal swabs, a daily stool sample (using provided at home collection kit) and skin swabs. In the intervals between sampling time points, the following data will be collected: vital signs, anthropometrics, all study self-reported measures (measures will be completed once during the four visits, and will be split into batches to prevent fatigue), short physical performance battery, leg press testing, submaximal bicycle testing (Astrand-Rhyming test, also serves as familiarization for maximal exercise test) and spirometry (Figure 3).

**Figure 3.**
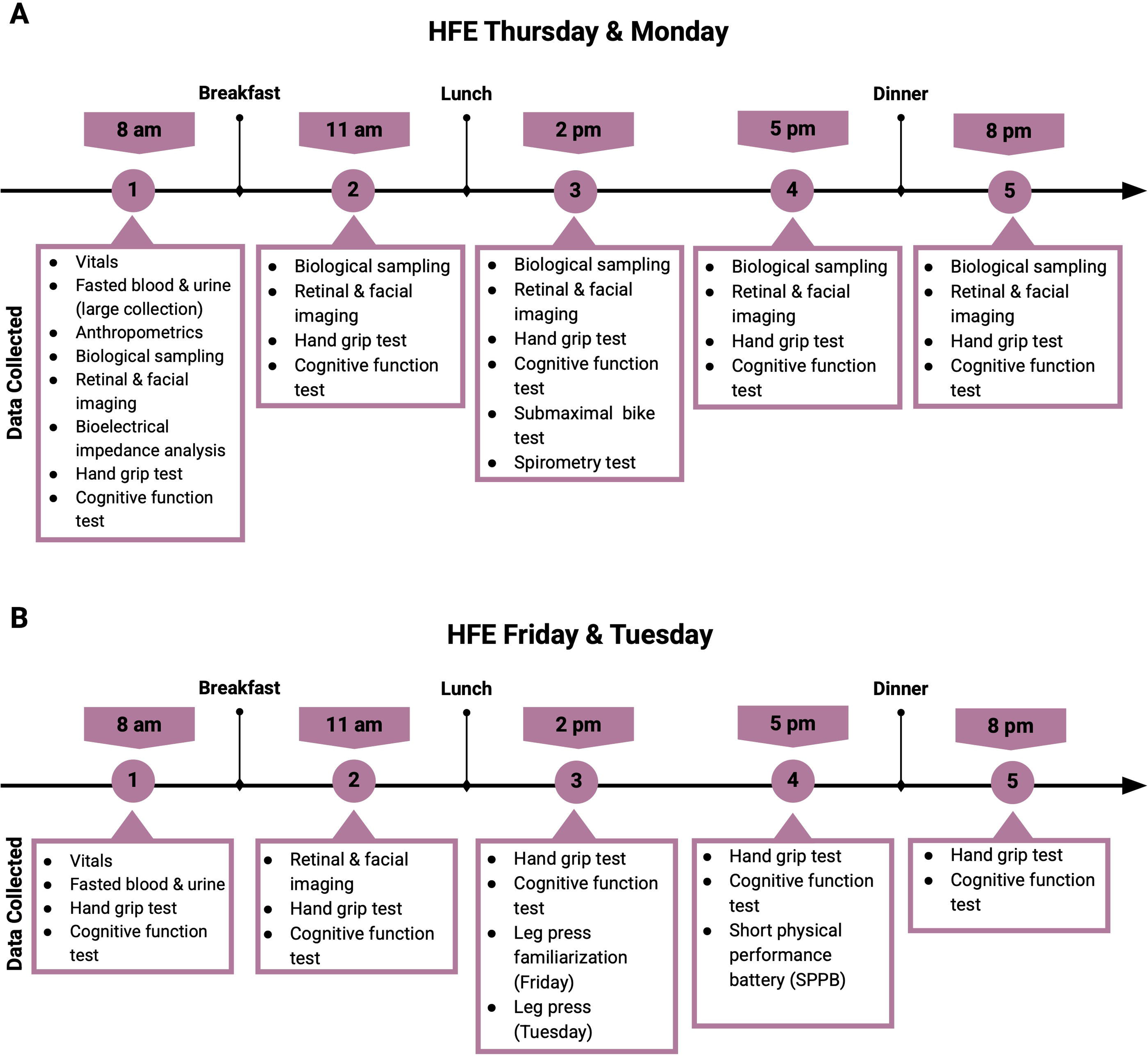
HFE sampling block. A) HFE sampling schedule on Thursday and Monday. B) HFE sampling schedule on Friday and Tuesday.

Three Study Meals will be provided at ∼0845H (immediately following the first data collection point), and during windows centered at 1200H and 1800H; or as desired by the participant. Photographs taken after each meal will be used to assess compliance and intake. Snacks, water, tea and coffee will be provided ad libitum, and all items consumed, and time of consumption will be recorded. Subjects will be asked to remain in the clinic and limit ambulatory activity.

At home between HFE visits, subjects will collect saliva samples before bed and on waking and collect a pooled overnight urine sample (Thursday and Monday). All meals over the weekend between the HFE visits will be provided by the study team and the following recommendations will be made for the weekend days between HFE visits: follow their usual weekend exercise and social patterns, and avoid extreme deviations (e.g., exercise longer or harder than usual, social gatherings that lead to extreme change their alcohol consumption or sleep schedule). Data will be captured over the weekend from the study wearables, allowing for an accounting of the effects of behaviors on biomarker data.

#### 2.7.7 Challenge Visit: Maximal Exercise Test (between weeks 6 – 10)

At the start of the visit, an IV cannula will be inserted and the LFE fasted blood sample will be collected following completion of other LFE measures. Then a standard meal will be provided and participants will rest for at least 1h before the exercise test. Immediately prior to the test, EKG leads are placed for live reading during the test. Maximal aerobic capacity will be assessed via a graded exercise test to volitional exhaustion on a Lode cycle ergometer. Participants will wear a mouthpiece with nose clip for continuous breath-by-breath gas analysis (ParvoMedics TrueOne 2400). Workload will begin at 5–50 W and increase by 3–30 W·min^-1^ according to fitness level, aiming for test duration of 8–12 min after warm-up. Continuous measurements include oxygen uptake (VO□), carbon dioxide production (VCO□), minute ventilation (VE), respiratory exchange ratio (RER), heart rate (ECG or heart rate strap), and blood pressure at 2-min intervals. Perceived exertion is recorded using the Borg scal^44^. Tests are terminated upon volitional fatigue, inability to maintain cadence, or meeting standard clinical termination criteria. Maximal effort will be confirmed by RER ≥ 1.10, heart rate within 10 bpm of age-predicted maximum, and/or a VO₂ plateau (<0.2 L·min^-1^ change over final 2 min). Venous blood samples will be collected at 0, 15, 30, 60, 90 and 120 (+/-5) minutes after the end of the test, and cognitive testing occurs and 30 minutes post-test (Figure 4).

**Figure 4.**
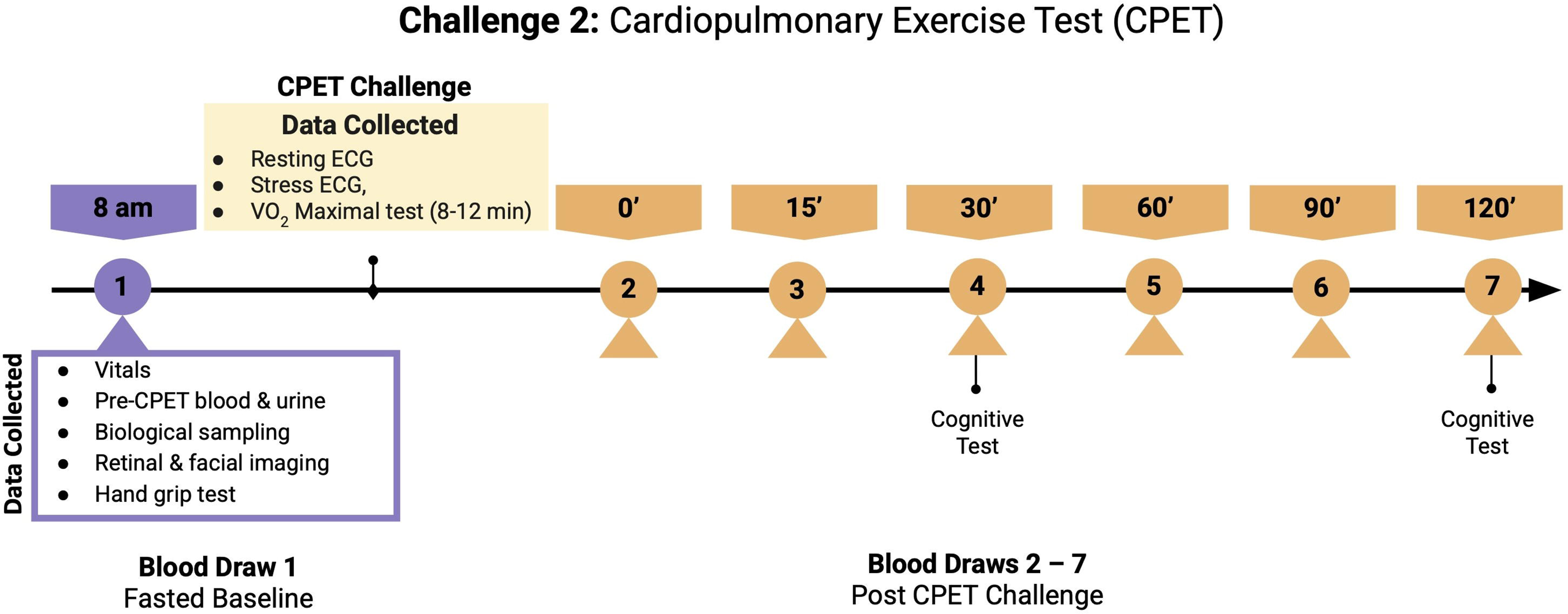
Challenge 2: Cardiopulmonary Exercise Test (CPET) Sampling Schedule

### 2.8 Follow-up Phase (6 and 12 months)

Follow-up in person visits replicate the structure of the Main Phase LFE visits. Vital signs and basic anthropometric data will be captured, along with blood for multi-omics analysis and clinical labs will be collected. Biological samples (stool, blood, urine, saliva, buccal swab, skin swab, vaginal swab) will be collected, grip strength will be tested, a retinal scan and facial images captured, and participants will complete a subset of study self reported measures and a subset of the cognitive tests.

## 3 Statistical Analysis

### 3.1 Power Calculation

A proprietary dataset^23^ containing multi-omic data similar to what is being collected in this study was used to calculate the required sample size for gleaning meaningful information from the multi-omic data collected as part of the TIME Study. The inter and intra-subject means and standard deviations of multi-omics from this dataset, which had measured >2000 multi-omic biomarkers, were used to randomly sample, representing the biological variation. The synthetic proteomic and metabolomic data were further scaled according to the coefficients of variation provided for each biomarker from Olink and Metabolon respectively, to represent technical variation. An effect size of 0.4795 was introduced to 10% of omics for half of the subjects. Power analysis was performed using ‘TTestIndPower’ from python’s statsmodels library^45^, with a desired power of 80% and an alpha of 0.05. A Bonferroni-corrected significance level was used to account for 12 effective dimensions resulting from performing a weighted gene co-expression network analysis (WGCNA)^46^, adjusting the alpha level accordingly. 100 iterations yielded a sample size requirement of 121 to detect an effect size bigger than 0.48 with a power of 80%. Notably, additional power will be gained through techniques such as feature pre-filtering and the use of regularization within ML algorithms. Thus, the enrollment target for the TIME Study was set as up to n =150, allowing for the contingency of not all individuals fully completing the study from start to finish, thereby meeting the calculated n = 121 individuals.

### 3.2 Analysis Plan

The statistical analysis plan for the study is designed to characterize the dynamic interplay within the human phenome by integrating various data modalities that collectively capture as many aspects of human physiology as possible. One of the primary objectives is to bridge the gap between discrete molecular data and near-continuous digital health data. Thus, we will process and analyze five primary data modalities: molecular data (including -omic data such as proteomics, metabolomics, lipidomics, and transcriptomics), digital data (near-continuous data collected from digital health technologies, including the Oura Ring, WHOOP Band, and continuous glucose monitor), clinical data (standard clinical laboratory measurements, anthropometric data, and vital signs), environmental data (publicly available data on air and water quality, and land cover), and ground truth labels (provided by participants via self report to identify specific activities).

The analysis will focus on four main areas: basic analysis for comparison to known; validated clinical measures; pattern discovery at the individual level; pattern discovery from aggregated population data; and the refinement of novel technologies for health and disease monitoring. Initially, all data will be processed and cleaned into a unified schema using established pipelines to ensure data quality and consistency by addressing missing data, outliers, and batch effects. Descriptive statistics will then be computed to summarize the data and identify preliminary patterns. All data will be ingested into and stored in a centralized cloud repository with a unified data schema built on a knowledge graph. This structure is crucial for integrating the disparate data types and applying various machine learning and artificial intelligence tools.

Using these multimodal data, we will define a temporal hierarchy of phenomic signal^47^, map molecular signals to digital health technology signals, refine the correlation structure of molecular signals in response to perturbations, and identify repeated cyclic modes within the data structure. We will apply a variety of dynamical systems analysis methods, such as time-scale decomposition, spectral analysis, and neural networks, to understand the temporal dynamics across the various data modalities. This approach will be complemented by the integration of the multimodal data described above using statistical analysis and machine learning techniques to merge the disparate data streams. Further modeling will then identify relationships and predict future states through both supervised and unsupervised machine learning. We will employ hypothesis testing with appropriate statistical tests to validate specific relationships between the data modalities. Missing data will be imputed using various metrics as previously described^23,24^.

In the future, we hope to analyze the samples from study meals to investigate research questions relating to precision nutrition. Such analyses would link the molecular profile of consumed food (the input) to the subsequent molecular changes in human biospecimens (the output). This framework would involve a three-step process. First, input profiling captures a complete molecular picture of the dietary input using a multi-omic approach on both single ingredients and the final, prepared meal. Second, output profiling measures the human biological response by analyzing the molecular composition of biospecimens (e.g., blood, urine, stool). This step includes establishing a preprandial baseline in a fasted state, followed by collecting postprandial samples at multiple time points to capture the dynamic processes of digestion, absorption, and metabolism. Finally, data integration and biomarker discovery would link the input and output data to identify food-specific molecular signatures that reliably appear in biofluids post-consumption.

## 4 Data Management and Dissemination

Data will be entered into a secure, web-based electronic case report form (eCRF, RedCap) during study visits by study staff. Each participant will be assigned a unique study ID, and all identifying information will be stored separately from study data. Data will be checked for completeness and range validity, and pipelines will address missing data, outliers, and batch effects to ensure data quality. Access to study data will be restricted to the Principal Investigator and authorized study personnel, and all data will be stored on encrypted servers at The Buck Institute. Every effort will be made to maintain subject confidentiality before, during, and after the study by implementing a coding system and removing identifiable information.

Where possible, we aim to share data with participants to incentivize retention. Participants can access their own wearable data throughout the study, and will be provided with a summary report at the end of the main phase. We will share hard copies of participant’s clinical laboratory results and results of the body composition testing and the maximal exercise testing. As the multiomic data are not actionable or clinically validated, we do not plan to share this data.

We are committed to the sharing of final research data, being mindful that the rights and privacy of people who participate in research must be protected at all times and that there is the need to protect patentable and other proprietary data. All datasets will be free of any identifiers that would permit linkages to individual research participants and variables that could lead to deductive disclosure of individual participants. Detailed data management procedures, including SOPs for data entry, coding, security, and storage, are available upon request. Anonymized raw, subject-level data will be held at The Buck Institute under the custody of the Principal Investigator and de-identified data will be made available upon reasonable written request to the Principal Investigator. The results of the study will be disseminated via publication in scientific papers, abstracts, and presentations. Study results will be posted in trial registries (e.g., WHO, ClinicalTrials.gov) and summarized in plain-language summaries accessible to the public. Authorship will be determined by ICMJE guidelines, manuscripts will be prepared by the study team.

## 5 Discussion

Characterizing the temporal dynamics of a living system is crucial to understanding how the system works. Human biology is incredibly complex, with many processes operating at different timescales. Here, we describe a study targeted at quantifying these temporal dynamics in humans across data modalities ranging from DHTs to blood proteins and metabolites to the gut and vaginal microbiomes and functional measures—the TIME Study. The design of the TIME Study has several important implications for human research.

First, TIME will serve as a reference dataset that quantifies biological variability across both daily (circadian/ultradian) and weekly timescales in healthy older adults. The structure intentionally mirrors real-world longitudinal multi-omics studies to ensure relevance and scalability, including allowance for naturalistic variation in sampling times and seasonal effects. We anticipate that the resulting dataset will facilitate the development of novel statistical models for the robust characterization and adjustment of time-of-day and day-of-week effects on thousands of biological features. This foundational data—comprising on the order of 10,000 unique biological and digital signals at each time point—will directly inform the optimization of future longitudinal study designs by empirically defining the necessary temporal resolution and sampling frequency required to capture specific, highly dynamic biological signals.

Second, the inclusion of controlled physiological perturbations (an OGTT and a CPET) allows for the study of these same signals in response to acute metabolic and physiologic stress, and is critical for resolving the dynamic regulatory capacity of the biological system. Studying this stress response is akin to assessing a car’s engine under load (the perturbation) rather than merely observing it while idling (the baseline state). The engine’s performance metrics (molecular and physiological outputs) following a rapid acceleration or hill climb (the challenge) reveal its true functional limits and efficiency—specifically, how quickly and effectively the biological machinery returns to a stable state (homeostasis). These perturbations are necessary to study the system’s resilience and adaptive speed, providing a far deeper measure of health than static baseline monitoring.

Third, TIME will help drive the biomedical community toward the definition of a Minimum Viable Dataset (MVD). The MVD is defined as the smallest set of measurements and corresponding optimal sampling frequency required to capture essential phenomic variation. Such a definition would be crucial for reducing costs and improving scalability across the biomedical pipeline. In drug discovery, an MVD could streamline early-stage target validation by ensuring that molecular screening accounts for the known, significant temporal variability of the target protein or metabolite. For pharmacological studies, it could define the minimum necessary data to accurately model pharmacokinetics and pharmacodynamics, thereby optimizing dosing schedules to align with a patient’s natural rhythms. Critically, in large-scale clinical trials, an MVD would provide a standardized, resource-optimized framework, ensuring that essential molecular and digital markers are collected with sufficient fidelity to minimize spurious results, maximize the statistical power required for personalized health interventions, and minimize the cost of such studies. While different applications will undoubtedly have different MVD requirements (e.g., a study investigating Alzheimer’s Disease will likely need to measure different biomarkers than a study investigating Type 2 Diabetes), TIME will provide unique insights into and interpretation of the temporal aspects of when to be measuring certain molecular and digital quantities.

The chosen study design introduces several documented and controlled constraints. Specifically, the protocol does not implement systematic control for seasonal variation across the recruitment and sampling periods. This is mitigated by the comprehensive documentation of all sampling dates and times, which supports post-hoc statistical modeling to characterize and adjust for these effects. Furthermore, the study utilizes single-site, regional recruitment spanning the Pacific and Mountain time zones in the USA. While this approach simplifies multi-site data harmonization, it limits the immediate generalizability of the findings to the specific recruited population. As the study has an extremely high participant burden due to intensive visit schedules, the population who consent to participate will unavoidably select for currently unknown lifestyle or background characteristics, although we manage this through flexible scheduling and enhanced engagement strategies.

Ultimately, the successful execution of the TIME Study will establish a critical baseline reference for the dynamic human phenome. It will provide insights into the significant operational logistics required to temporally profile the human system, lowering the activation energy required to run future studies on other cohorts that might study important biological patterns such as the female menstrual cycle. Finally, it will help define a MVD, centered around DHT combined with specimen-derived biomarkers, that can inform the design of future biomedical studies. Once complete, the TIME resource stands to significantly advance our capacity to investigate the crucial role of time in homeostasis, health, and age-related decline.

## Supporting information

TIME Study SPIRIT Figure

TIME SPIRIT 2025 checklist

## Data Availability

No data associated with this work

## Abbreviations

AHA: American Heart Association
AI: Artificial Intelligence
CGM: Continuous glucose monitor
CPET: Cardiopulmonary Exercise test
CRU: Clinical Research Unit
DHTs: Digital Health Technologies
ECG: Electrocardiogram
eCRF: Electronic case report form
HFE: High frequency epoch
ICMJE: International Committee of Medical Journal Editors
IRB: Institutional Review Board
IV: intravenous
LFE: Low frequency Epoch
MVD: Minimum viable dataset
OGTT: Oral Glucose Tolerance test
PBMC: Peripheral blood mononuclear cell
PI: Principal investigator
RER: Respiratory exchange ratio
SMS: Short message service
SPIRIT: Standard Protocol Items Recommendations for Interventional trials
TIME: Temporal Investigation of Multimodal Elements
VE: Minute ventilation
VO2max: Maximal oxygen uptake
WHO: World Health Organization

The study protocol and any subsequent amendments were reviewed and approved by the Advarra Institutional Review Board (IRB reference:Pro00085349), 6100 Merriweather Dr., Suite 600, Columbia, MD 21044, call toll free; 877-992-4724). The TIME study is registered with ClinicalTrials.gov (NCT07107386, https://clinicaltrials.gov/study/NCT07107386). Trial registration date: 2025-06-15. All participants provided written informed consent prior to enrollment and participation in any study procedures.

## 7 Author Contributions

Conceptualization, [J.T.Y].; methodology, [J.T.Y, B.J.S]; writing—original draft preparation, [J.T.Y, E.G, B.J.S]; writing—review and editing [all]; project administration, [J.T.Y, B.J.S]; supervision: [J.T.Y, B.J.S]; funding acquisition, [J.T.Y, B.J.S]; All authors have read and agreed to the published version of the manuscript.

## 10 Acknowledgements

The authors sincerely thank the study participants for their time and commitment.

## 11 Consent for publication

Not applicable. No individual identifying data or images are included in this manuscript.

## 12 Availability of data

De-identified data, including a data dictionary and, where feasible, statistical code, will be available to qualified researchers upon reasonable written request to the Principal Investigator, with review to ensure participant privacy and protection of proprietary information.

