## Supplementary material for "The Temporal Investigation of Multimodal Elements (TIME) Study: Protocol for an observational, longitudinal study to characterize the dynamic structure of molecular and digital data in healthy older adults": TIME Study SPIRIT Figure

**Figure 1:** Full study overview in SPIRIT figure format.

|  | **Screening** | | **Main 11-week study period**  ***(visits may take place over 15 weeks if needed)*** | | | | | **Follow Up period** |
| --- | --- | --- | --- | --- | --- | --- | --- | --- |
| **Visit Name** | **Phone Screen** | **Initial Visit** | **Low Frequency Epoch (LFE) Visits** | **At home procedures** | **LFE + Oral Glucose Challenge (Additional procedures)** | **High Frequency Epoch Visits** | **LFE+ Maximal Exercise Challenge (Additional procedures)** | **Follow Up Visits** |
| **Timeline** | **Up to 30 days before Initial Visit** | **Up to 30 days before LFE1** | **Week 1; 9 weekly visits until Week 11 *(+4 weeks)*** | **Daily, Week 1 – 12 months** | **Once, between Week 2 - 5** | **4 days, (Th, Fri, Mon, Tues) Between Week 4 and Week 8** | **Once, between Week 7 -10** | **6 and 12 months *(+/- 2 weeks)*** |
| Study information | X | X |  |  |  |  |  |  |
| Eligibility assessment | X | X |  |  |  |  |  |  |
| Written consent |  | X |  |  |  |  |  |  |
| Health history (and updates) |  | X | X |  |  | X |  | X |
| Medication and supplement use (and updates) |  | X | X |  |  | X |  | X |
| Demographics |  | X |  |  |  |  |  |  |
| Anthropometrics |  |  | X (V1 and V9) |  |  | X |  | X |
| Vital signs |  | X | X |  |  | X |  | X |
| EKG |  | X |  |  |  |  | X |  |
| Venous blood sample |  |  | X |  | X | X | X | X |
| Buccal Swab |  |  | X |  | X | X | X | X |
| Saliva sample |  |  | X |  | X | X | X | X |
| Vaginal swab |  |  | X |  | X | X | X | X |
| Skin Swab |  |  | X |  | X | X | X | X |
| Fasted urine sample |  |  | X |  | X | X | X | X |
| Urine pooled collection |  |  |  |  |  | X |  |  |
| Stool sample |  |  | X |  | X | X | X | X |
| Retinal Fundus Imaging |  |  | X |  | X | X | X | X |
| Facial Imaging |  |  | X |  | X | X | X | X |
| Study self-reported measures |  |  | X (V1 and V9) |  |  | X |  | X |
| Study meals |  |  |  |  | X | X | X |  |
| Study wearables/devices |  |  | X | X | X | X | X | X |
| Fasting Resting Metabolic Rate |  |  | X (V1and V9) |  |  |  |  |  |
| Cognitive testing |  |  | X |  | X | X | X | X |
| Grip Strength Testing |  |  | X |  | X | X | X | X |
| Short Physical Performance Battery |  |  |  |  |  | X |  |  |
| Leg press familiarization |  |  |  |  |  | X |  |  |
| Leg press testing |  |  |  |  |  | X |  |  |
| Submaximal bicycle exercise |  |  |  |  |  | X |  |  |
| Spirometry |  |  |  |  |  | X |  |  |
| Oral glucose tolerance test |  |  |  |  | X |  |  |  |
| Maximal bicycle exercise test |  |  |  |  |  |  | X |  |
